# Time series forecasting of COVID-19 confirmed cases with ARIMA model in the South East Asian countries of India and Thailand: a comparative case study

**DOI:** 10.1101/2020.05.16.20103895

**Authors:** Viswa Chaitanya Chandu

## Abstract

**Background:** As economic burden makes it increasingly difficult for countries to continue imposing control measures, it is vital for countries to make predictions using time series forecasting before making decisions on lifting the restrictions.

**Aim:** Since apparent differences were noted in the disease transmission between the two South East Asian countries of India and Thailand, the study aims to draw comparative account of the progression of COVID 19 in near future between these two countries.

**Methods:** The study used data of COVID 19 confirmed cases in India and Thailand from WHO COVID 19 situation reports during the time period between 25^th^ March, 2020 and 14^th^ May, 2020. After determination of stationarity in the data and differencing, observation of autocorrelation function (ACF) and partial autocorrelation function (PACF), Auto Regressive Integrated Moving Average (ARIMA) (2,2,1) model was used to forecast the COVID 19 confirmed cases in both these countries for two weeks (i.e. 28^th^ May, 2020). IBM SPSS version 20.0 software was used for data analysis.

**Results:** The study demonstrated a possible increasing trend in number of COVID 19 cases in India in the coming two weeks with an estimated point forecast of 1,28,772 (95% CI 115023–142520) by 28^th^ May, 2020. A stationary phase was forecasted for Thailand with a difference of only 43 cases between 14^th^ May (the last case of input data) and 28^th^ May.

**Conclusion:** The time series forecasting employed in the present study warrants thorough preparation on part of the Indian health care system and authorities and calls for caution with regard to decisions made on lifting the control measures. The difference in the time series forecasting between these two South East Asian countries also highlights the need for strengthening of public health systems.

## Introduction

Coronavirus disease (COVID-19) is one of the greatest challenges the world has encountered in recent times. Since the initial reports of outbreak in late December, 2019, the numbers have been consistently rising with the disease affecting 4.3 million people in 181 countries worldwide as of 15^th^ May, 2020.^1^ The frailty of a multitude of health care systems across the globe has been exposed by COVID-19. With the surfacing negative socioeconomic consequences of non-pharmaceutical interventions like lockdown affecting the vulnerable, especially in developing countries, and the plethora of uncertainties around eradication of COVID-19, governments are eyeing at easing the restrictions that have long been in place.^2^ It is imperative to understand that lifting the control measures for economic salvage, without thoroughly preparing for the possible consequences, may only result in further economic decline and health crisis. WHO in its strategic advice for countries looking to life the control measures illustrated six criteria in a sequential manner to be considered: control of transmission; preparation of health systems for active contact tracing and optimum care provision; careful management of health facilities to prevent outbreaks; adherence to preventive measures as the essential services resume; management of importation risks; indoctrination of the ‘new norm’ among communities by active engagement.^3^ In this scenario, it is vital for countries to make predictions using time series forecasting. Since apparent differences were noted in the disease transmission between the two South East Asian countries of India and Thailand, the study aims to draw comparative account of the progression of COVID-19 in near future between these two countries.

## Methods

The time series analysis in this study was based on the daily number of laboratory confirmed cases reported from 25^th^ March, 2020 to 14^th^ May, 2020 collected from the WHO COVID-19 situation reports.^4^ In this study Auto Regressive Integrated Moving Average (ARIMA) model, an advanced time series forecasting technique was employed. ARIMA models decompose the past data basing on the knowledge of past values (autoregressive component), stabilizing the data pattern (integrated component), and adjusting for the past error terms (moving average component), which are expressed as p, d, q respectively.^5^ The data from both the countries were analyzed in the following manner: a) inspection for stationarity using sequence charts and correlograms; b) differencing to transform non-stationary data to stationary; c) creation of ARIMA models based on the autocorrelation function (ACF) and partial autocorrelation functions (PACF); d) determination of ARIMA(p,d,q) model fit; e) forecasting the time series for next two weeks i.e. till 28^th^ May, 2020; f) cross validation of the ARIMA model by comparison of predicted confirmed cases with the true confirmed cases over the time period from which the data was collected. IBM SPSS Version 20.0 software (IBM SPSS statistics for windows version 20, Armonk, NY, USA) was used for data analysis.

## Results

Preliminary observation of data from both the countries revealed an upward quadratic trend. Since the data pattern did not demonstrate stationarity with first order differencing, second order differencing was done to achieve stationarity. Figure 1 shows the second order differenced data pattern for both the countries showing stationarity. On observation of ACF and PACF plots with second order differencing, a single significant spike in ACF was observed at Lag 1 and two spikes were observed in PACF at Lag 1 and lag 2, which were suggestive of MA(1) and AR(2), respectively (Figure 2). Based on these observations, ARIMA(2,2,1) modeling was performed. The model demonstrated good fit (Table 1). Figure 3 shows the ACF and PACF of the residuals of ARIMA (2,2,1). The number of confirmed cases was forecasted for a period of two weeks in both the countries (Table 2, Figure 4). Figure 5 shows the cross validation of the performed ARIMA model by comparing the model predicted confirmed cases with true confirmed cases from 25^th^ March, 2020 to 14^th^ May, 2020. The high correlation between the predicted and true values highlights the predictive accuracy of the model. Another important observation from this study is that the time series forecasting based on past data predict a stationary phase in Thailand, where as the trend in India was predicted to be considerably rising. According to the results from ARIMA(2,2,1) forecasting in the present study, the confirmed cases in India by 28^th^ May, 2020 has a point forecast of 1,28,772 (95% CI 1,15,023 – 1,42,520).

**Figure 1:**
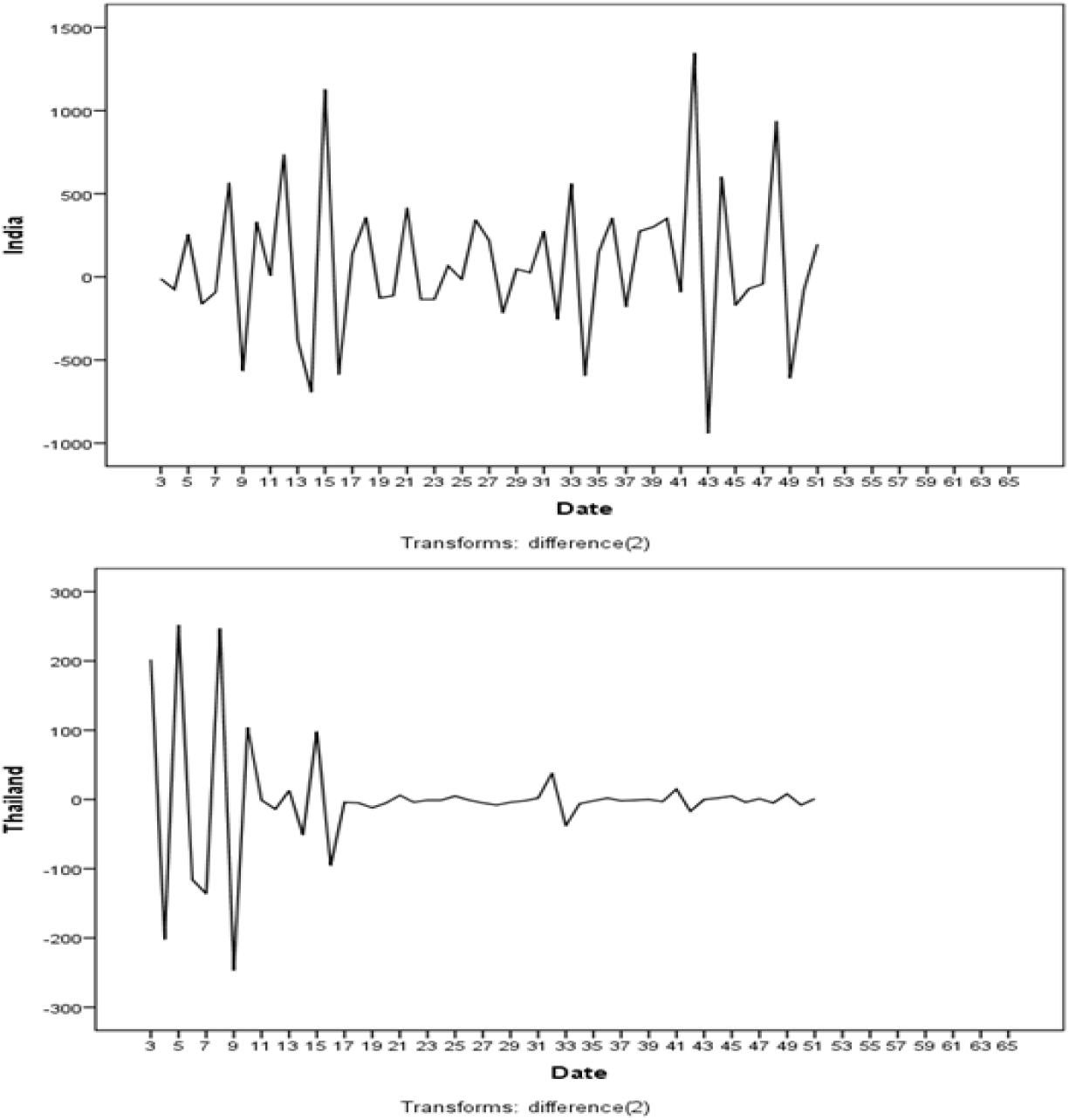
Second order differenced sequence chart of both the countries demonstrating stationarity of data pattern.

**Figure 2:**
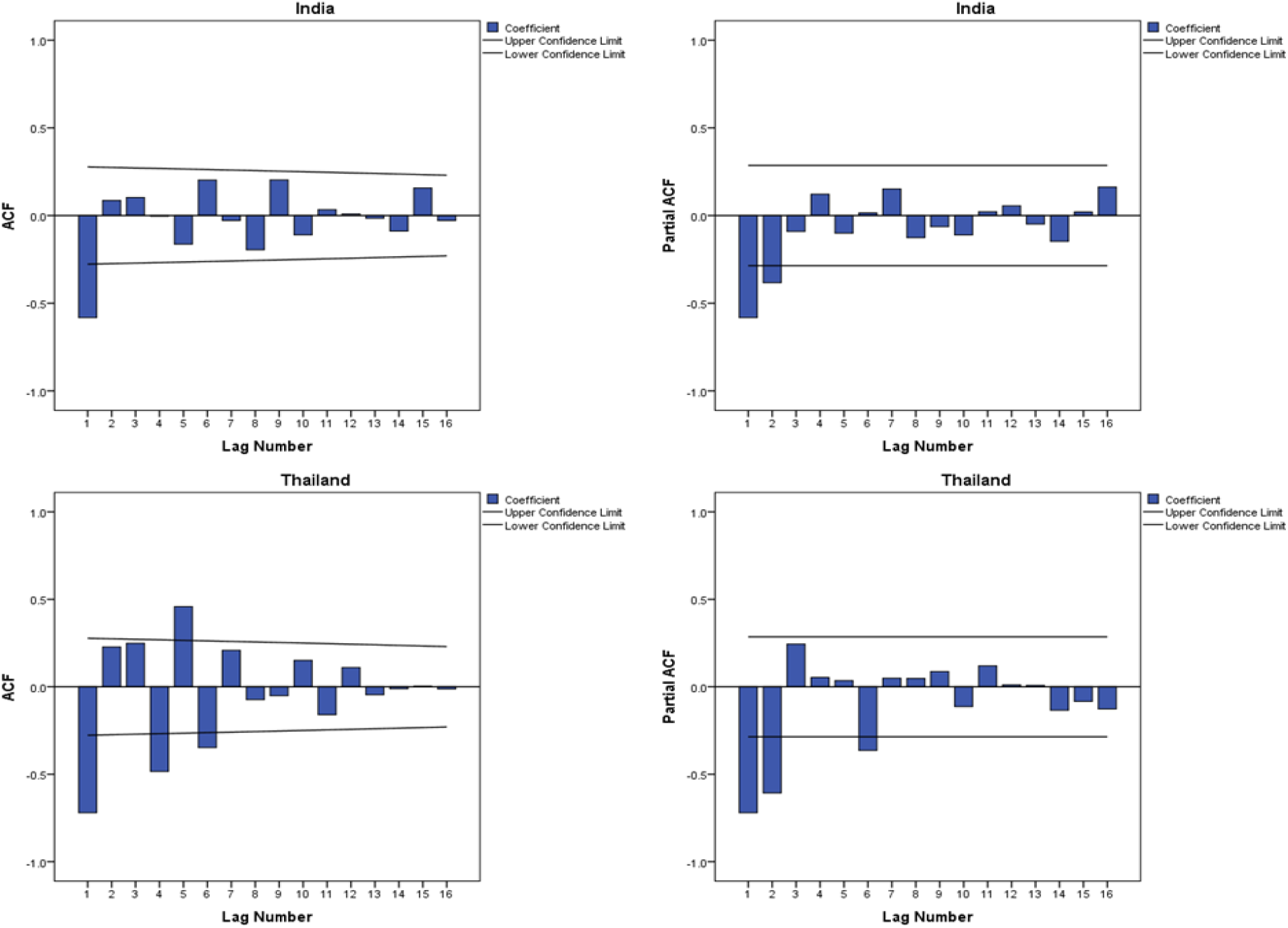
Second order differenced ACF and PACF plots.

**Figure 3:**
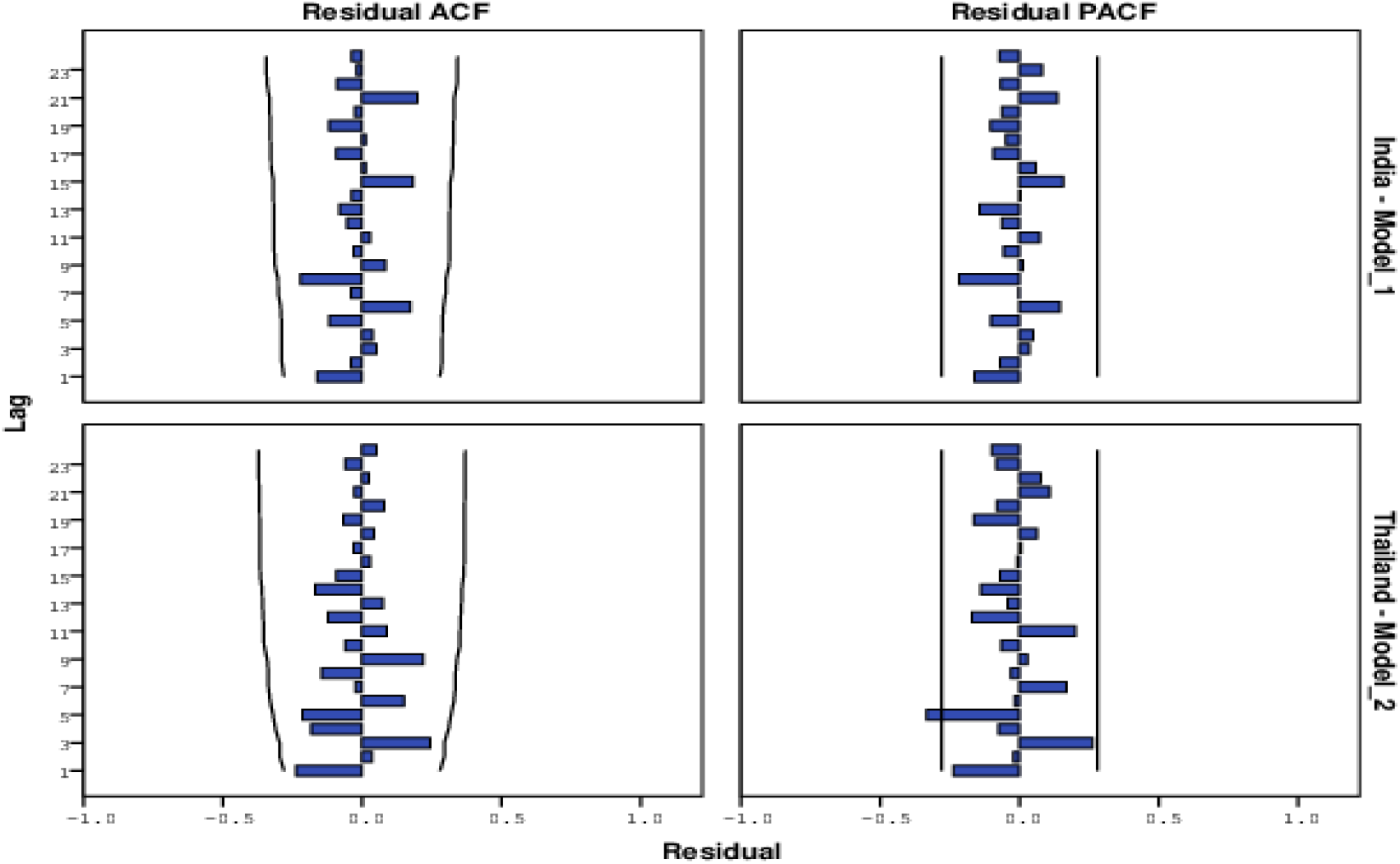
ACF and PACF of residuals of ARIMA(2,2,1)

**Figure 4:**
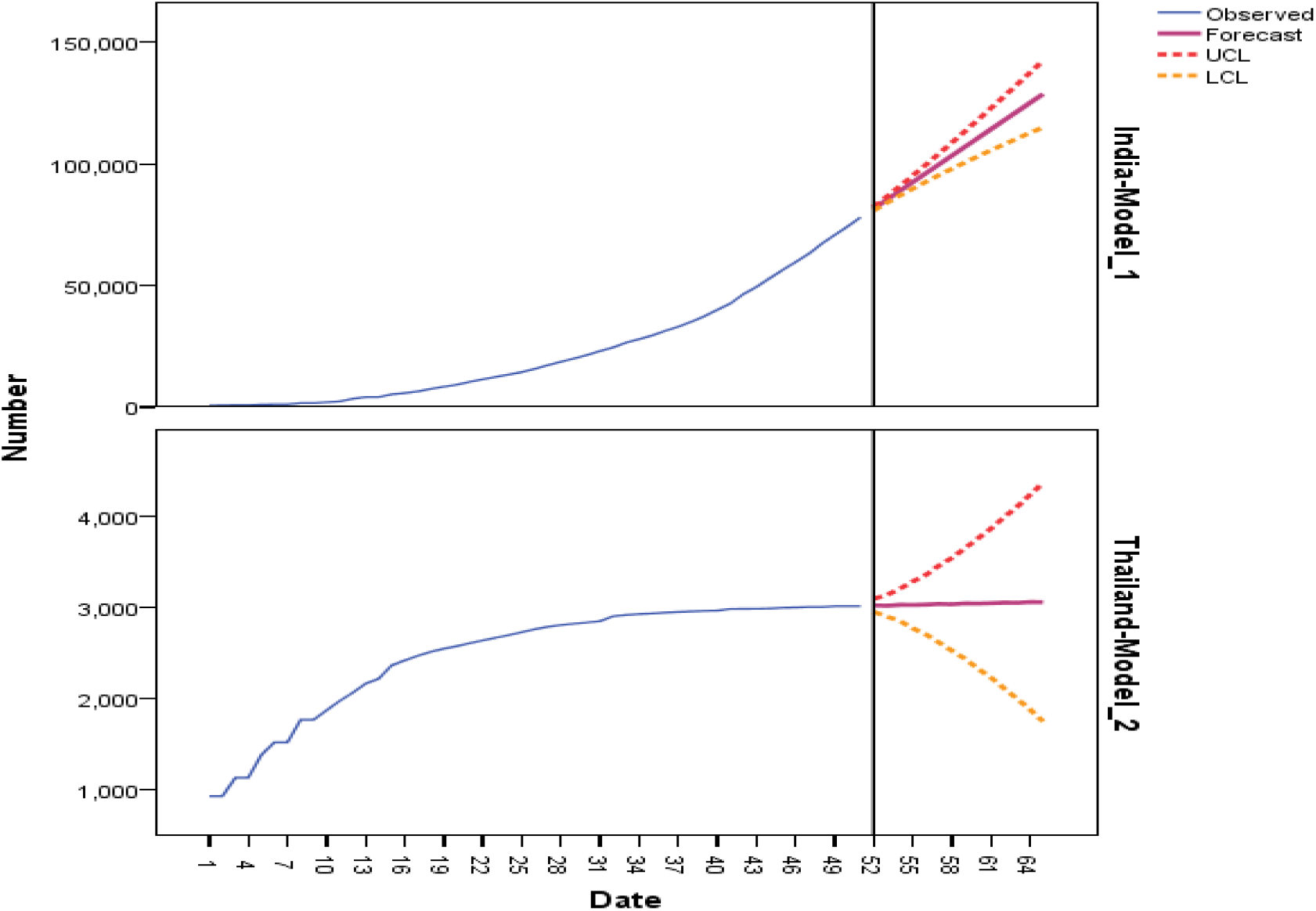
Forecasting of COVID-19 confirmed cases in India and Thailand (till 28^th^ May, 2020)

**Figure 5:**
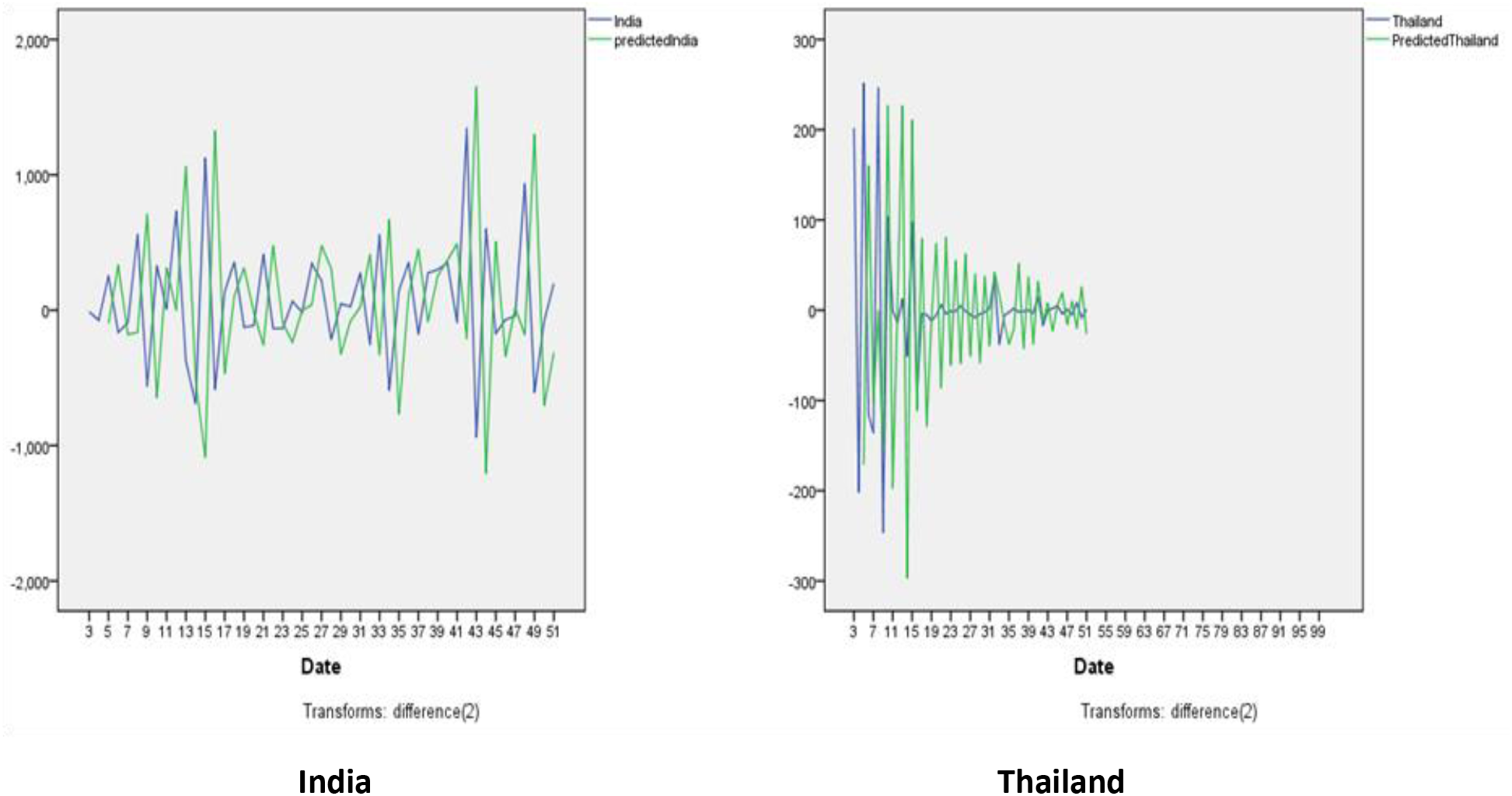
Cross validation of ARIMA(2,2,1) model by comparing model prediction and true data (25^th^ Match, 2020 to 14^th^ May, 2020)

**Table 1:** ARIMA(2,2,1)* Model Parameters Country.

| Country | Parameter | Value | Standard Error | T statistic | Ljung Box Q(18) sig. | R<sup>2</sup> |
| --- | --- | --- | --- | --- | --- | --- |
| India | AR(2) | -0.317 | 0.314 | -1.008 | 0.698<sup>†</sup> | 1.00 |
|  | MA(1) | -0.135 | 0.601 | -0.224 |  |  |
| Thailand | AR(2) | -0.926 | 0.04 | -20.26 | 0.125<sup>†</sup> | 0.993 |
|  | MA(1) | -0.913 | 0.136 | -6.702 |  |  |

\*constant was not considered due to second order differencing; <sup>†</sup> non significance; R<sup>2</sup> coefficient of determination

**Table 2:** Forecasting of COVID-19 confirmed cases in India and Thailand with point forecast and 95% confidence intervals (till 28^th^ May, 2020)

| INDIA |  |  | THAILAND |  |  |
| --- | --- | --- | --- | --- | --- |
| Date | Point forecast | 95% confidence interval | Date | Point forecast | 95% confidence interval |
| 15<sup>th</sup> May | **81590** | 80832 - 82349 | 15<sup>th</sup> May | **3027** | 2955 – 3099 |
| 16<sup>th</sup> May | **85222** | 83954 – 86491 | 16<sup>th</sup> May | **3022** | 2904 – 3141 |
| 17<sup>th</sup> May | **88862** | 86949 – 90775 | 17<sup>th</sup> May | **3033** | 2857 – 3208 |
| 18<sup>th</sup> May | **92481** | 89798 – 95164 | 18<sup>th</sup> May | **3031** | 2773 – 3290 |
| 19<sup>th</sup> May | **96114** | 92610 – 99618 | 19<sup>th</sup> May | **3035** | 2709 – 3360 |
| 20<sup>th</sup> May | **99742** | 95333 – 104151 | 20<sup>th</sup> May | **3041** | 2621 – 3462 |
| 21<sup>st</sup> May | **103370** | 97990 – 108750 | 21<sup>st</sup> May | **3038** | 2530 – 3547 |
| 22<sup>nd</sup> May | **107000** | 100588 – 113411 | 22<sup>nd</sup> May | **3048** | 2442 – 3654 |
| 23<sup>rd</sup> May | **110628** | 103125 – 118131 | 23<sup>rd</sup> May | **3046** | 2332 – 3761 |
| 24<sup>th</sup> May | **114257** | 105606 – 122907 | 24<sup>th</sup> May | **3051** | 2234 – 3868 |
| 25<sup>th</sup> May | **117885** | 108035 – 127736 | 25<sup>th</sup> May | **3056** | 2118 – 3994 |
| 26<sup>th</sup> May | **121514** | 110412 – 132616 | 26<sup>th</sup> May | **3055** | 2001 – 4108 |
| 27<sup>th</sup> May | **125143** | 112741 – 137544 | 27<sup>th</sup> May | **3063** | 1885 – 4241 |
| 28<sup>th</sup> May | **128772** | 115023 - 142520 | 28<sup>th</sup> May | **3061** | 1753 - 4370 |

## Discussion

The key reason behind comparison of Indian and Thai contexts was to verify if stronger public health systems, matched for geographical and cultural background to a major extent, can play a role in successfully fighting the COVID-19 global pandemic. A comparative account of few indicators comparing the Thai and Indian health care system is provided in Table 3. From a broader perspective, the distribution of health professionals at different operational levels was considerably equitable in Thailand regardless of the economic status of the provinces, which could be a result of the focused health infrastructure development since the past few decades. Concentration on improving the primary health-care system encompassing optimum number of health care workers at different operational levels and increasing the financial risk protection, to ensure financial hardships do not come in the way of accessing quality health care, have been the most prolific measures adopted by Thailand which made the country set an example for developing world.^6^ To compare a few health system related parameters between India and Thailand, the latter offers health care delivery predominantly through Ministry of Public Health (MOPH) facilities while the former’s health care delivery is increasingly been privatized. The appointments of all the health professionals in Thailand are regular, while India has preferred contractual employment.^7^ Thailand has special tracks to enroll students from underserved areas and rural communities into medical schools since 1974^8^; India does not have such mechanisms. The comparison of nurse and midwifery professionals per 10,000 population and community health workers between Thailand (27.59 and 10,64,434 respectively) and India (17.271 and 9,70,676 respectively) reveals a huge disparity at these operational levels between the health systems.^9^ Moreover, Thailand spends 3.208% of GDP as public health expenditure; whereas India’s public spending on health care is 1.4% of GDP.^10^ While it is impossible to ignore the huge difference in the size of populations between these countries, there are obvious lessons that Indian health care system can emulate from Thai’s, which include: strengthening public health care delivery systems; restructuring the medical education system in an informed manner; preparing health work force at various levels; increasing public health expenditure to at least 3% GDP.

## Conclusion

The numbers from the time series forecasting employed in the present study warrant thorough preparation on part of the Indian health care system and authorities and call for caution with regard to decisions made on lifting the lockdown. While Thailand is preparing for easing the lockdown restrictions from 17^th^ May, 2020, it is apparent that a fourth lockdown may be considered in India after 17^th^ May. While COVID-19 certainly has been an unfortunate pandemic of unprecedented magnitude in recent times, it also poses questions which many systems around the world have conveniently ignored over the years, and thus provides an important opportunity for the health systems to revive and restructure.

## Data Availability

The data analyzed in this study is available.

